# *ABCB1* GENE POLYMORPHISMS ARE ASSOCIATED WITH CLINICAL RESPONSE TO NABIXIMOLS IN PATIENTS WITH MULTIPLE SCLEROSIS-RELATED SPASTICITY

**DOI:** 10.1101/2025.06.20.25329925

**Authors:** Alessandra Gemma, Marco Mauri, Paola Banfi, Maurizio Versino, Alen Zollo, Filippo Martinelli Boneschi, Franca Marino, Marco Cosentino, Marco Ferrari

## Abstract

**Background:** Multiple Sclerosis (MS) is an autoimmune disease of the central nervous system causing severe symptoms, including spasticity-related pain. Nabiximols (NBX), a drug combining Tetrahydrocannabinol and Cannabidiol, is approved for MS-associated spasticity. Despite NBX efficacy, only 60-70% of patients respond to treatment. This study explores whether genetic polymorphisms in key genes involved in NBX pharmacology influence response to drug.

**Materials & Methods:** This is a genetic, exploratory study in which MS patients, treated with NBX, were retrospectively enrolled. Polymorphisms in genes related to NBX metabolism (*CYP2C9*, *CYP2C19*), efflux (ATP Binding Cassette subfamily B member 1 - *ABCB1*), and cannabinoid receptors (*CNR1*, *CNR2*) were analyzed using Real-Time PCR.

**Results:** Among 45 enrolled patients, 29 responded to NBX and 16 did not. Of responders, 38% were homozygous for the T allele in ABCB1 1236C>T (rs1128503), and 48% for the T allele in ABCB1 3435A>T (rs1045642). None of the non-responders carried T allele. The odds ratios for response to NBX of TT homozygotes was 20.5 (95% CI:1.1-376.1, P=0.0039) for rs1128503 and 30.9 (95% CI:1.7-563.2, P=0.0006) for rs1045642.

**Discussion:** This study suggests a link between genetic profiles and NBX response. If validated in larger studies, these findings could pave the way for personalized therapy for MS-related spasticity.

## Introduction

Multiple Sclerosis (MS) is a chronic inflammatory, demyelinating, and neurodegenerative disease affecting the central nervous system [1]. MS patients experience a wide range of symptoms, with spasticity affecting at least two-thirds of individuals [2]. Nabiximols (NBX), oromucosal spray (Sativex®), drug containing Tetrahydrocannabinol (THC) and cannabidiol (CBD), is approved for adult patients with moderate-to-severe MS-associated spasticity [3]. Despite its efficacy, NBX elicits a treatment response in only 60–70% of MS patients [4], and currently, no reliable predictors of its effectiveness are available for use in clinical practice. Genetic factors represent crucial contributors to variability in drug response. In particular, single nucleotide polymorphisms (SNPs) may predict drug efficacy and safety in many clinical settings [5].

Both THC and CBD act on cannabinoid receptors (CBR) 1 and 2, which are encoded by the cannabinoid receptor genes (*CNR*) 1 and 2 [6]. SNPs in *CNR1*, such as rs1049353 and rs2023239 and in *CNR2* (rs2501431), influence various receptor functions, including depression [7], happiness perception [8] and childhood obesity [9]. THC and CBD are substrates for P-glycoprotein (P-gp), an efflux pump encoded by the ATP Binding Cassette Subfamily B Member 1 (*ABCB1*) gene [6]. SNPs in the *ABCB1*, such as rs1128503 and rs1045642, have been associated with interindividual variability in drug response [10], including response to cannabinoids [11]. SNPs in the *ABCB1* have also been proposed as potential predictors of interindividual variability in response to opioids [12] as well as cannabis-based therapies [13]. Additionally, THC and CBD are metabolized by the CYP450 enzyme family, coded by *CYP2C9*, *CYP2C19*, and *CYP3A4* genes [6,11], and it has been shown that SNPs, such as rs1799853 (*CYP2C9*) and rs4244285 (*CYP2C19*), dramatically reduce metabolic activity of these enzymes [14].

In the present study, we investigated the relationship between all abovementioned SNPs and response to drug treatment in a cohort of MS patients in which MS-related spasticity was treated with NBX.

## Methods

### Patients

This exploratory genetic study retrospectively enrolled all patients diagnosed with MS based on the McDonald criteria 2017 and treated with NBX for MS-related spasticity at the "Centro Sclerosi Multipla, Ambulatorio Malattie Demielinizzanti - Ospedale di Circolo e Fondazione Macchi, Varese, Italy" from May 2014 to June 2024. Exclusion criteria were: i) use of other spasticity-related pain medications, such as anticonvulsants/antiepileptic, antidepressants, or opioids, ii) presence of kidney or liver diseases; iii) concurrent treatments with drugs known to affect NBX metabolism or transport; iv) other conditions/disease causing chronic pain (e.g., cancer, trigeminal neuralgia, fibromyalgia, diabetes, arthritis).

The study received approval from the Ethics Committee of ASST Sette Laghi, Varese, Italy, on February 21, 2023 (Study number 164). Informed consent and blood samples for genotyping were collected from all participants during routine follow-up visits from May 2023 to June 2024.

Response to NBX was evaluated using the Patient-rated spasticity 0-10 Numeric Rating Scale (NRS) as well as through clinical evaluation [15–16]. According to the current accepted definition, a reduction of ≥ 20% from baseline on the NRS after 4 weeks of treatment represents a minimal clinically difference, while a reduction of ≥ 30% indicates a clinically important difference [15–16].

On this basis, we decided to consider patients who achieved at least a 30% reduction in NRS at week 4 compared to baseline and continued with NBX treatment, as responders.

Patients who did not achieve a 30% reduction in NRS scores within the fourth week of treatment and were switched to alternative spasticity therapy were considered as non-responders.

### SNPs selection criteria and genotyping

SNPs in *CNR*1 (rs1049353 and rs2023239); *CNR*2 (rs2501431); *ABCB1 (*rs1128503 and rs1045642); *CYP2C9* (rs1799853) and *CYP2C19* (rs4244285), were selected based on their frequency in the Caucasian population, which must be greater than 10%, and the availability of information regarding their biological and clinical effects. A detailed description of SNPs included in the study are reported in **Table 1**.

**Table 1.** SNPs included in the study.

| Gene | Variant | N.C. | A.F. (%) | Biological effect |
| --- | --- | --- | --- | --- |
| *CNR1* | rs1049353 | 1359G>A | 27 | Associated to addiction [23] and happiness [8]. |
|  | rs2023239 | -3163A>G | 17 | Major risk of adverse effects [24]. |
| *CNR2* | rs2501431 | 24201643G>A | 58 | Major risk of depression [7]. |
| *ABCB1* | rs1128503 | 1236C>T | 43 | Lower expression [17]. |
|  | rs1045642 | 3435A>T | 52 | Lower expression [25, 17]. |
| *CYP2C9* | rs1799853 | 9133C>T | 12 | Reduced activity [14] |
| *CYP2C19* | rs4244285 | 681G>A | 15 | No activity [14]. |

Abbreviations: N.C., nucleotide change; A.F., allelic frequency in Caucasian population; *ABCB1*, ATP
Binding Cassette subfamily B member 1; *CYP*, CYtochrome P450; *CNR1*, CaNnabinoid Receptor 1 gene;
*CNR2*, CaNnabinoid Receptor 2 gene.

DNA was extracted using FTA Elute Cards (GE Healthcare Bio-Sciences AB, SE-751 84 Uppsala, Sweden) according to the manufacturer’s instructions (https://it.vwr.com/store/product/7997552/fta-elute-cards-whatmantm).

Selected SNPs were genotyped using a Real-Time PCR system (StepOne®, Thermo Fisher Scientific, Waltham, MA, USA) with a pre-designed TaqMan® genotyping assay (Thermo Fisher Scientific, Waltham, MA, USA). SNPs were analyzed using 100 ng of genomic DNA in a 25 μl reaction containing 12.5 μl of TaqMan Universal Master Mix and 1.25 μl of each specific TaqMan probe (Thermo Fisher Scientific, Waltham, MA, USA). PCR amplification was carried out under the following thermal cycling conditions: an initial activation step at 95°C for 10 minutes, followed by 40 cycles of denaturation at 95°C for 15 seconds and annealing/extension at 60°C for 60 seconds. A final read step was performed at 60°C for 30 seconds.

### Statistical analysis

Data are shown as the mean ± standard deviation (SD), unless otherwise stated. The statistical significance of the differences between groups was assessed by the Mann–Whitney U-test or by One-way analysis of variance followed by Bonferroni’s Multiple Comparison Test as appropriate. The evaluation of Hardy-Weinberg equilibrium was assessed using the χ2-test (P<0.05). Differences in allele frequencies between groups were analyzed by the χ2-test for trend or the Fisher’s exact test using a recessive model (wild type/heterozygous vs homozygous for SNP). The odds ratio (OR) with a 95% confidence interval (CI) was calculated using a recessive model.

Statistical analyses were performed using GraphPad Prism version 5.00 for Windows (GraphPad Software, San Diego, California, USA, www.graphpad.com).

## Results

### Patients

From clinical records, we identified 47 patients treated with NBX for MS-related spasticity from May 2014 to June 2024. Of these, one patient was excluded due to concomitant opioid treatment for cancer-associated pain, and another was excluded for using carbamazepine to manage trigeminal neuralgia-related pain. **Table 2** shows demographic and clinical characteristics of the 45 patients finally enrolled.

**Table 2.** Clinical and demographic characteristics of MS patients. * = P<0.001 vs non-responders.

|  | All | Responders | Non-responders |
| --- | --- | --- | --- |
| Number of subjects | 45 | 29 | 16 |
| Gender (male/female) | 18/27 | 14/15 | 4/12 |
| Age (years, mean±SD) | 53,0±11,1 | 53,1±12,2 | 52,9±9,2 |
| Disease duration (years, mean±SD) | 17,3±10,1 | 16,3±10,6 | 19,0±9,2 |
| EDSS (mean±SD) | 5,6±1,8 | 5,4±1,8 | 5,9±1,6 |
| MSSS (mean±SD) | 6,2±2,1 | 6,1±2,2 | 6,2±2,0 |
| **MS type** |  |  |  |
| RR | 25 | 19 | 6 |
| PP | 11 | 6 | 5 |
| SP | 9 | 4 | 5 |
| **NBX dosage** (puffs/dye, mean±SD)* | 5.6±2.5 | 5.8±1.8 | 5.4±3.0 |
| **NRS score** |  |  |  |
| before NBX (mean±SD) | 7,1±1,2 | 7,1±1,4 | 7,1±1,2 |
| after NBX (mean±SD) | 4,1±2,1 | 2,6±1,8* | 5,9±0,8 |
| % reduction (mean±SD) | 47,0±28,5 | 64,7±21,1* | 14,7±2,0 |

EDSS, Expanded Disability Status Scale; MSSS, Multiple Sclerosis severity scale; MS, multiple sclerosis;
RR, relapsing-remitting; PP, primary progressive; SP, secondary progressive; THC, Δ9-tetrahydrocannabinol; CBD, cannabidiol; NRS, numerical rating scale. \*, 1 puff NBX = 100 µl spray including 2,7 mg THC and 2,5 mg CBD.

Among the enrolled patients, 29 achieved an NRS score reduction of 30% or greater after 4 weeks of NBX treatment and were included in the responder group. For 16 patients, the NRS score reduction did not reach 30%. These patients switched to alternative pain treatments and were included in the non-responder group.

We did not find any difference between groups in terms of gender, age, MS type, disease duration, MS therapy, Multiple Sclerosis Severity Score (MSSS), Expanded Disability Status Scale (EDSS), or NBX dosage (**Table 2**).

### Correlation between patient genotype and response to NBX

All SNPs were in Hardy–Weinberg equilibrium (data not shown). Among the 29 patients who responded to NBX therapy, 18 (62%) were carriers of the C allele in the rs1128503 SNP in *ABCB1*, and 15 (52%) were carriers of the A allele in the rs1045642 SNP in *ABCB1*, while 11 (38%) and 14 (48%), respectively, were homozygous for the T allele. None of the 16 patients who did not respond to NBX therapy were homozygous of the T allele (**Table 3**). Using a χ2 test for trend, we found that the frequency of the T allele in both rs1128503 (1236C>T) and rs1045642 (3435A>T) in *ABCB1* was significantly higher in responders compared to non-responders (P < 0.0010 and P < 0.0012 respectively). This result was confirmed by the Fisher exact test. The odds ratio (O.R.) for response to NBX was 20.5 (95% C.I.: 1.1–376.1, P = 0.0039) for rs1128503, and 30.9 (95% C.I.: 1.7–563.2, P = 0.0006) for rs1045642. SNPs in *CNR*1, *CNR*2, *CYP2C9*, and *CYP2C19* were not significantly associated with the response to NBX (**Table 3**).

**Table 3.**
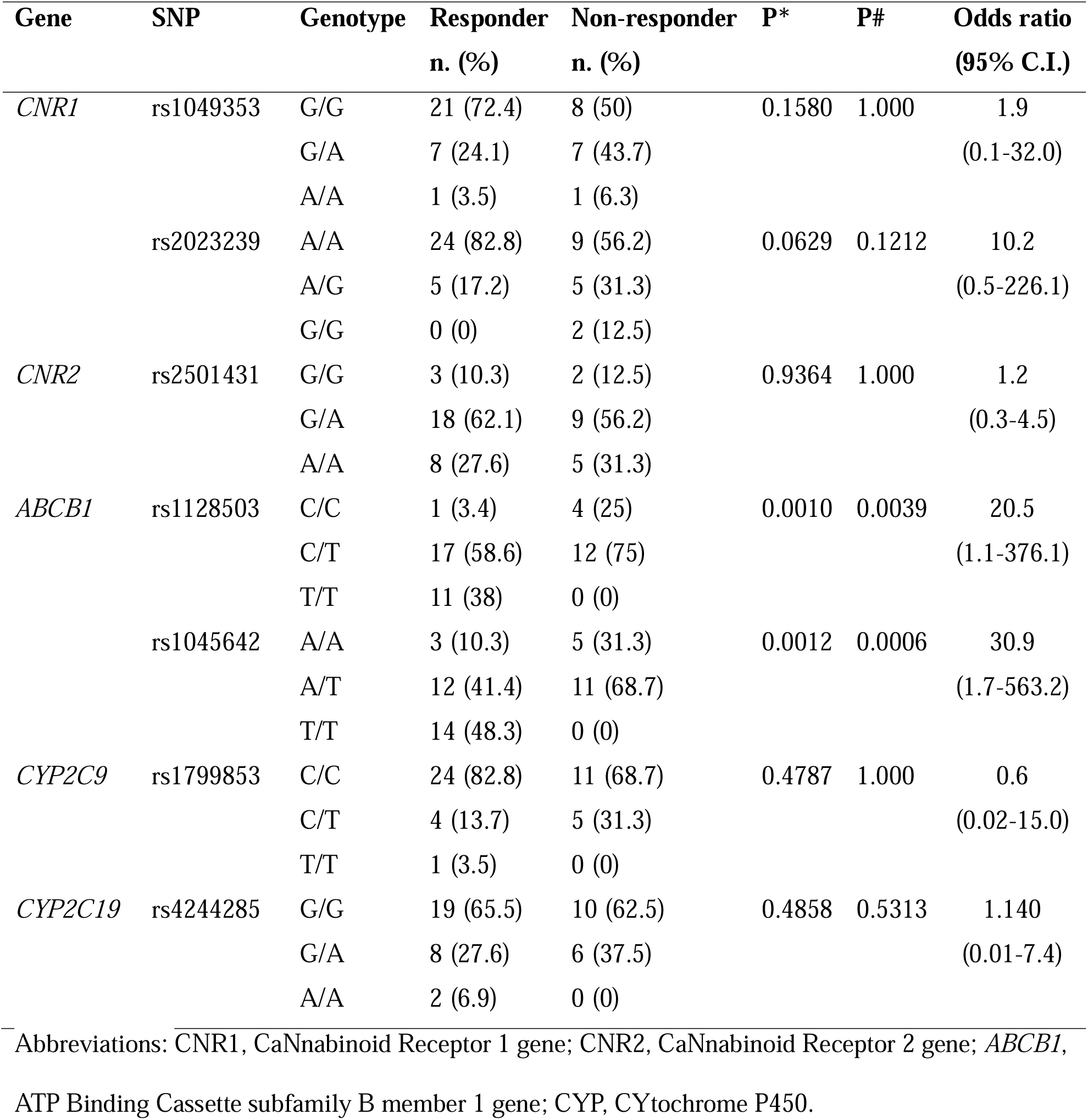
Correlations between patient’s genotype and NBX response. * = χ2-test for trend; # = Fisher Exact Test.

| Gene | SNP | Genotype | Responder<br>n. (%) | Non-responder<br>n. (%) | P* | P# | Odds ratio<br>(95% C.I.) |
| --- | --- | --- | --- | --- | --- | --- | --- |
| *CNR1* | rs1049353 | G/G | 21 (72.4) | 8 (50) | 0.1580 | 1.000 | 1.9 |
|  |  | G/A | 7 (24.1) | 7 (43.7) |  |  | (0.1-32.0) |
|  |  | A/A | 1 (3.5) | 1 (6.3) |  |  |  |
|  | rs2023239 | A/A | 24 (82.8) | 9 (56.2) | 0.0629 | 0.1212 | 10.2 |
|  |  | A/G | 5 (17.2) | 5 (31.3) |  |  | (0.5-226.1) |
|  |  | G/G | 0 (0) | 2 (12.5) |  |  |  |
| *CNR2* | rs2501431 | G/G | 3 (10.3) | 2 (12.5) | 0.9364 | 1.000 | 1.2 |
|  |  | G/A | 18 (62.1) | 9 (56.2) |  |  | (0.3-4.5) |
|  |  | A/A | 8 (27.6) | 5 (31.3) |  |  |  |
| *ABCB1* | rs1128503 | C/C | 1 (3.4) | 4 (25) | 0.0010 | 0.0039 | 20.5 |
|  |  | C/T | 17 (58.6) | 12 (75) |  |  | (1.1-376.1) |
|  |  | T/T | 11 (38) | 0 (0) |  |  |  |
|  | rs1045642 | A/A | 3 (10.3) | 5 (31.3) | 0.0012 | 0.0006 | 30.9 |
|  |  | A/T | 12 (41.4) | 11 (68.7) |  |  | (1.7-563.2) |
|  |  | T/T | 14 (48.3) | 0 (0) |  |  |  |
| *CYP2C9* | rs1799853 | C/C | 24 (82.8) | 11 (68.7) | 0.4787 | 1.000 | 0.6 |
|  |  | C/T | 4 (13.7) | 5 (31.3) |  |  | (0.02-15.0) |
|  |  | T/T | 1 (3.5) | 0 (0) |  |  |  |
| *CYP2C19* | rs4244285 | G/G | 19 (65.5) | 10 (62.5) | 0.4858 | 0.5313 | 1.140 |
|  |  | G/A | 8 (27.6) | 6 (37.5) |  |  | (0.01-7.4) |
|  |  | A/A | 2 (6.9) | 0 (0) |  |  |  |

Abbreviations: CNR1, CaNnabinoid Receptor 1 gene; CNR2, CaNnabinoid Receptor 2 gene; *ABCB1*, ATP Binding Cassette subfamily B member 1 gene; CYP, CYtochrome P450.

Subjects with the T/T and C/T genotypes for the rs1128503 SNP in *ABCB1* exhibited a significantly greater percentage reduction in NRS scores following NBX treatment compared to those with the ancestral C/C genotype (P < 0.05 and P < 0.001 respectively). Similarly, patients with the T/T genotype for the rs1045642 SNP showed a significantly greater percentage reduction in NRS scores compared to both the C/C and C/T genotypes (P < 0.0001) (**Figure 1**). On the contrary SNPs in the *CNR* and *CYP* don’t showed any notable association with NRS scores (**data not shown**).

**Figure 1.**
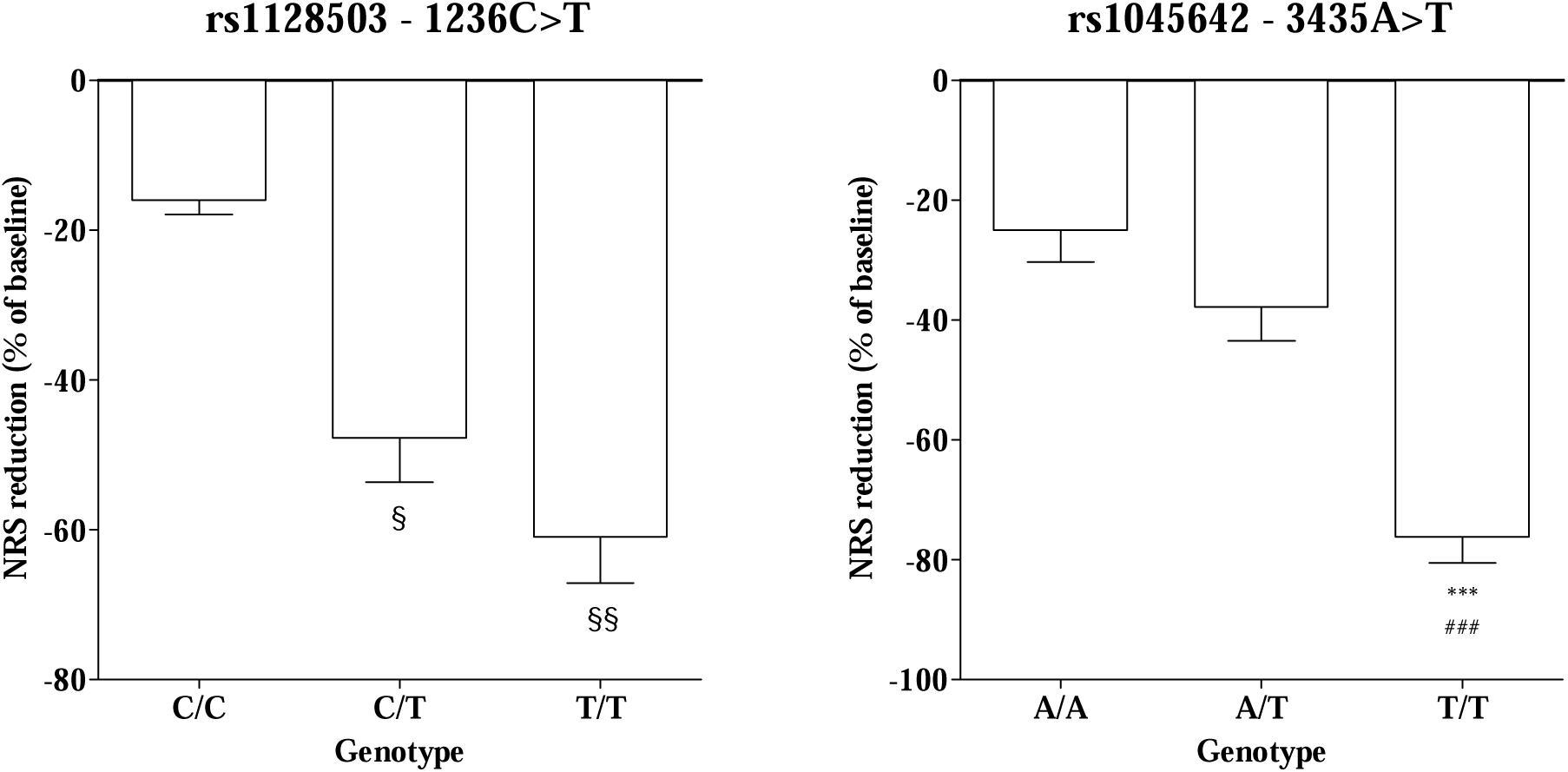
Relationship between SNPs *ABCB1* and % of reduction in NRS score. § = P < 0.05 vs C/C; §§ = P < 0.01 vs C/C; *** = P < 0.0001 vs A/A; ### = P < 0.0001 vs A/T.

## Discussion

The main finding of this study is that the T allele in the *ABCB1* SNPs rs1128503 (1236C>T) and rs1045642 (3435A>T) is predictive of response to NBX treatment for MS-related spasticity.

Specifically, the T allele, previously associated with reduced P-glycoprotein (P-gp) efflux activity [17], was more frequent among treatment responders. Moreover, patients carrying the T allele in either SNPs showed significantly greater percentage reductions in NRS scores following NBX treatment compared to those with ancestral or heterozygous genotypes.

To date, only a limited number of studies have evaluated the role of SNPs in *ABCB1* in response to cannabinoid treatments. Among these, Poli and colleagues [13] demonstrated that haplotypes containing the rs1045642 SNP may influence the effectiveness of cannabis in managing chronic pain, as well as its associated side effects. Beyond *ABCB1* SNPs, other SNPs have also been linked to cannabinoid response. For instance, the *CYP2C9*\*2 and *CYP2C9*\*3 SNPs were associated with higher plasma levels of CBD [18] and THC-induced side effects [19], moreover, in epilepsy patients, SNPs in *ABC* family gene were found to enhance CBD response [20]. Finally, SNPs in the *CNR1* gene have been shown to affect THC’s impact in patients with irritable bowel syndrome [21].

In light of these findings, several factors, including SNPs in *ABCB1*, *CYP2C9*, and *CNR*1, have been proposed as potential determinants of cannabinoid-related dependence and side effects [22].

However, so far, reliable indicators to predict the efficacy of NBX in clinical practice have to be identified yet.

In this study, we found, for the first time, that patients carrying the T allele in the *ABCB1* SNPs rs1128503 and rs1045642 exhibited a better response to NBX. Our hypothesis is that higher frequencies of SNPs known to reduce P-gp activity may increase NBX bioavailability and facilitate its passage across the blood-brain barrier. This could result in higher drug concentrations at its site of action, thereby improving NBX efficacy. These findings hold promise for advancing precision medicine by helping to identify patients most likely to benefit from NBX treatment, potentially improving its efficacy by enhancing its presence at the site of action. However, further validation in larger patient cohorts is necessary before this method can be considered a reliable tool for predicting patient response to NBX therapy in clinical settings.

We acknowledge that our study has some limitations, primarily the retrospective design and the small sample size. However, it must be considered that this is an exploratory study with strict inclusion criteria (i.e. Analgesic monotherapy with NBX, exclusion of concomitant treatments with drugs influencing NBX metabolism/transport, and/or diseases causing chronic pain). The strict inclusion criteria reduce possible confounders and, in turn, increase the likelihood of evaluating the role of genetics in response to NBX treatment. Moreover, although the number of enrolled patients was relatively low, the allelic frequency of the selected SNPs is always above 10%, thus reducing the number of subjects needed to reach the study goals.

In conclusion, in this exploratory study, we have shown, for the first time, a relationship between a patient’s genetic profile and response to NBX treatment. If confirmed in a prospective study involving a larger cohort of patients, our results could pave the way for the identification of new, useful tools for predicting the response to NBX treatment in MS patients suffering from spasticity, ultimately allowing for personalized therapy in patients with indications for this drug.

## Ethical approval

All procedures performed in studies involving human participants were in accordance with the ethical standards of the institutional committee (ASST Sette Laghi Hospital, Varese, Italy on February 21, 2023 - Study number 164) and with the 1964 Helsinki declaration and its later amendments or comparable ethical standards. Informed consent was obtained from all individual participants included in the study prior to their inclusion in the study.

## Data Availability Statement

The data that support the findings of this study are available from the corresponding author upon reasonable request. Data are in controlled access data storage at Centre for Research in Medical Pharmacology, University of Insubria, Varese, Italy.

## Authorship contributions

MC, MV, MM, FMB, AG and MF conceptualized the manuscript. MM, PB, AZ and AG collected the clinical data. AG performed the genetic analysis of the patients. MF and AG drafted manuscript first version. MC, FM, MV, AG, AZ, FMB, MM, and MF participated in critical analysis and review of the manuscript. All authors read and approved the final manuscript.

## Data Availability

All data produced in the present study are available upon reasonable request to the authors

## Acknowledgements

The authors are grateful to Dr. Margherita Italia, Neurology and Stroke Unit, ASST Sette Laghi Hospital, Varese, Italy for her helpful collaboration in the collection and evaluation of clinical data. Alessandra Gemma developed a research program focused on innovative approaches to evaluating the role of polymorphisms in the *ABCB1* in clinical responses to nabiximols in patients with multiple sclerosis-related spasticity as part of her work for the PhD Course in Clinical and Experimental Medicine and Medical Humanities, University of Insubria (XXXVII Cycle).

## Additional information

The Authors wish to dedicate this study to the loving memory of Paola Banfi, dearest friend, caring physician, and invaluable scientist.

